# Lipoprotein(a): Particle vs. Mass and Cardiovascular Events in the Cardiovascular Risk in Young Finns Study population

**DOI:** 10.64898/2026.04.30.26352188

**Authors:** Olli Raitakari, Matias Knuuti, Anna Linko-Parvinen, Katja Pahkala, Suvi Rovio, Noora Kartiosuo, Sini Stenbacka, Irina Lisinen, Britt-Marie Loo, Terho Lehtimäki, Mika Kähönen, Claudia Lamina, Florian Kronenberg, Markus Juonala, Jorma S.A. Viikari, Juha Mykkänen

**Affiliations:** Centre for Population Health Research, University of Turku and Turku University Hospital, Turku, Finland; Research Centre of Applied and Preventive Cardiovascular Medicine, University of Turku, Turku, Finland; Department of Clinical Physiology and Nuclear Medicine, Turku University Hospital, Turku, Finland; InFLAMES Research Flagship, University of Turku, Turku, Finland; Clinical Chemistry, Tyks Laboratories, Turku University Hospital and University of Turku, Turku, Finland; Paavo Nurmi Centre & Sports and Exercise Medicine, University of Turku, Turku, Finland; Department of Public Health, University of Turku and Turku University Hospital, Turku, Finland; Department of Mathematics and Statistics, University of Turku, Turku, Finland; Joint Clinical Biochemistry Laboratory, Turku University Hospital, and University of Turku, Turku, Finland; Department of Clinical Chemistry, Fimlab Laboratories, and Finnish Cardiovascular Research Center - Tampere, Faculty of Medicine and Health Technology, Tampere University, Tampere, Finland; Department of Clinical Physiology, Tampere University Hospital, and Finnish Cardiovascular Research Center - Tampere, Faculty of Medicine and Health Technology, Tampere University, Tampere, Finland; Institute of Genetic Epidemiology, Medical University of Innsbruck, Innsbruck, Austria; Department of Medicine, University of Turku and Division of Medicine, Turku University Hospital, Turku, Finland

**Keywords:** Lipoprotein(a), Atherosclerosis, Coronary artery disease, Apolipoprotein(a), Genetic risk score

## Abstract

**Background:** Lipoprotein(a) [Lp(a)] can be reported as mass concentration (mg/dL) or particle concentration (nmol/L). Because Lp(a) biology is likely mediated at the particle level, molar concentration may better reflect biologically relevant exposure. We compared molar- and mass-based Lp(a) measurements in relation to cardiovascular outcomes, *LPA* genetic variation, and apo(a) isoform phenotype in a population-based cohort.

**Methods:** Lp(a) was measured in 6,182 participants in the Young Finns Study. Among participants aged ≥40 years, associations with prevalent coronary artery disease (CAD) and composite cardiovascular disease (CVD) were assessed using logistic regression models. Lp(a) was modeled as quintiles, inverse-normal transformed continuous variables, and clinically relevant cut-points. Discrimination and model fit were evaluated using c-statistics and Akaike’s Information Criterion. Associations with an *LPA* genetic risk score (GRS) and apo(a) isoform phenotype were examined using Spearman’s correlation analyses.

**Results:** The relation between molar and mass Lp(a) was not constant across the concentration range. The molar-to-mass ratio increased across higher clinically relevant Lp(a) categories, indicating concentration-dependent correspondence between nmol/L and mg/dL. The molar-to-mass ratio was directly associated with the *LPA* GRS and inversely associated with apo(a) isoform size, and these associations were more strongly attenuated after adjustment for molar than for mass Lp(a). Across CAD and composite CVD, molar- and mass-based Lp(a) showed broadly similar associations. In fully adjusted CAD models, the odds ratio for the highest versus lowest quintile was 1.55 for molar Lp(a) and 1.67 for mass Lp(a); corresponding continuous-model ORs were 1.20 and 1.22 per 1-unit increase in inverse-normal transformed Lp(a). Discrimination and global model fit were essentially identical between the two measurement scales.

**Conclusions:** Molar- and mass-based Lp(a) measurements showed comparable epidemiologic associations with prevalent cardiovascular outcomes. However, molar reporting aligned somewhat more closely with the genetic and structural determinants of Lp(a), supporting continued standardization toward particle-based reporting.

**Clinical Perspectives:** *What Is New?:*

- In the Young Finns Study, Lp(a) reported in molar and mass units showed highly similar associations with prevalent coronary artery disease and composite cardiovascular disease.
- The relation between molar and mass Lp(a) was not fixed: the molar-to-mass ratio varied across clinically relevant Lp(a) concentration ranges and according to apo(a) isoform size.
- Molar Lp(a) showed slightly stronger alignment with the *LPA* genetic risk score, and ratio-based analyses suggested that genetically and structurally determined assay discordance was more closely captured by molar than by mass measurement.

*What Are the Clinical Implications?:*

- Mass- and molar-based Lp(a) remain broadly comparable for cardiovascular risk association analyses, but they should not be treated as directly interchangeable using a single universal conversion factor.
- Particle-based reporting in nmol/L may better reflect the biologic dimension of Lp(a), even when disease discrimination is similar to that of mass-based reporting.

## Introduction

Lipoprotein(a) [Lp(a)] is a genetically determined plasma lipoprotein that has emerged as a causal risk factor for atherosclerotic cardiovascular disease (CVD). Large cohort studies and Mendelian randomization analyses have demonstrated a continuous association between elevated Lp(a) concentrations and coronary artery disease (CAD), ischemic stroke, and calcific aortic valve disease^1^. Lp(a) levels are largely genetically determined by variation in the *LPA* gene and remain relatively stable for decades from childhood to adulthood across the life course^2^, and predict adult cardiovascular outcomes, underscoring the long-term clinical relevance of early-life Lp(a) levels^3^.

Structurally, Lp(a) consists of a low-density lipoprotein (LDL)–like particle covalently linked via a disulfide bond to apolipoprotein(a) [apo(a)]. Apo(a) is a highly polymorphic glycoprotein that contains multiple kringle domains, most notably a variable number of kringle IV type 2 (KIV-2) repeats^4^. The number of KIV-2 repeats in the *LPA* gene varies from approximately 2 to more than 40 copies between individuals. Each kringle domain contains ∼80–85 amino acids and has a molecular weight of about 9–10 kDa, resulting in apo(a) isoforms comprising roughly 11 to over 50 kringle IV motifs and spanning molecular weights from ∼300 to 800 kDa^5^. When combined with the 2.7-3.1 MDa LDL-like moiety^6^, the molecular mass of a single Lp(a) particle varies by approximately 23-30% across the apo(a) isoform ranges.

This structural heterogeneity has important analytical implications. When Lp(a) measurement is based on and calibrated against mass concentration, the measurement reflects the total mass of particles present, which is a function of both particle number and molecular weight. Consequently, mass concentration does not directly reflect particle number. Because atherothrombotic activity is mediated at the particle level, it is presumed that cardiovascular risk relates more directly to particle number than to total particle mass^7^. For this reason, several expert groups and laboratory standardization initiatives have advocated measuring Lp(a) in molar units^1,8^. Nonetheless, mass-based measurement is common in clinical practice and in historical epidemiological cohorts. Because there is no fixed conversion factor between mg/dL and nmol/L, these two measuring systems are not interchangeable at the individual level^9^.

Thus, from a biological perspective, it is plausible that cardiovascular risk relates more directly to particle number than to total particle mass. However, comparative outcome data are limited. In an analysis of the ODYSSEY OUTCOMES trial, Lp(a) concentrations measured by mass- and molar-based immunoassay demonstrated nearly identical associations with major adverse cardiovascular events^10^. Whether this apparent equivalence extends to general population cohorts remains uncertain.

Therefore, in the present study, using data from the three-generational Young Finns Study (YFS) ^11^, we examined the association of Lp(a) with prevalent atherosclerotic cardiovascular events when measured as (1) particle concentration (nmol/L) and (2) mass concentration (mg/dL).

## Methods

The YFS is a prospective multicenter study from Finland. The first large baseline examination was conducted in 1980 (baseline age, 3–18 years, N=3,596)^12^. The latest follow-up in 2018-2020 was extended to cover the original participants as well as their parents and offspring aged 3 years and older^11^. After excluding those who had deceased, decided to withdraw from the study or for whom contact information was unavailable, we invited 3,217 original participants (Generation 1; aged 40-58 y). Additionally, we invited 5,696 offspring (Generation 2; 3-38 y), and 3,940 parents (Generation 0; 59-92 y) of the original participants, including parents and adult offspring of those original participants who had deceased. The details of the study design have been previously published^11^. Anonymized data are available upon reasonable request from the YFS research group (https://youngfinnsstudy.utu.fi/).

### Assessment of Lp(a) concentrations

Venous blood samples were drawn after at least a 4-hour fast. Separated plasma or serum was frozen in aliquots at -70°C. Lp(a) mass concentration was measured in year 2020, immediately after the completion of the latest field study^11^. Lp(a) molar concentration was measured from frozen samples in year 2025.

#### Lp(a) mass concentration

The mass concentration of Lp(a) was measured using an immunoturbidimetric method originally developed for Konelab instruments (Lipoprotein(a) system reagent, Thermo Fisher Scientific, Vantaa, Finland), which was modified for an Indiko Plus Clinical Chemistry Analyzer (Thermo Fisher Scientific, Vantaa, Finland). The assay was calibrated using manufacturer-provided calibrators derived from human serum, with concentrations assigned by the manufacturer. The overall interassay coefficient of variation for internal quality control samples was 6.2% at a nominal concentration of 12.5 mg/dL. The lower limit of quantification was 3.1 mg/dL.

#### Lp(a) molar concentration

Molar measurements of lipoprotein(a) were assessed with immunoturbidimetric method with Preciset Lp(a) Gen. 2 using a c702 module in a Cobas 8000 system using polyclonal antibodies raised in rabbits (Roche Diagnostics GmbH, Mannheim, Germany). The method is standardized against WHO/IFCC International Reference Reagent (SRM2B), a second-generation certified reference material used for assay calibration, which assigns its nanomolar values using ELISA methods based on monoclonal antibodies detecting a single epitope at KIV-9. Bilevel internal quality control samples were analyzed daily with mean (SD) 38.6 nmol/L (1.2) and 111.6 nmol/L (4.7), with overall interassay coefficient of variation of 3.6%. The lower limit of quantification was 7 nmol/L.

### Atherosclerotic cardiovascular outcomes

Linkages to national registries, including the Care Register for Health Care and the National Death Index, were used to ascertain outcomes, including CAD, atherosclerotic cerebrovascular disease and peripheral artery disease^3,13^. The outcomes included both thrombotic events and confirmed diagnoses without a thrombotic event. CVD events included the first instance of adjudicated myocardial infarction, stroke, transient ischemic attack, ischemic heart failure, CAD, peripheral artery disease, carotid intervention, abdominal aortic aneurysm, or coronary revascularization. All Finnish citizens are covered in the registry data.

The study protocol was approved by the appropriate institutional ethics committees, and all participants provided written informed consent in accordance with the Declaration of Helsinki.

### LPA genetic risk score

The whole genomes were genotyped using either an Illumina Infinium 670K or a Global Screening Array chip according to manufacturer protocols. Genotypes were imputed using Minimac3 and TOPMed r3 reference series on a TOPMed Imputation Server^14^. The *LPA* genetic risk score utilized in the current paper is adapted from a weighted genetic risk score described by Trinder *et al*.^15^, composed of 43 single-nucleotide polymorphisms, while 33 of these were found and harmonized across all three generations of the 2018 field study^15^. Due to skewness in its distribution, rank order normalization was applied when treating the LPA genetic risk score as a continuous variable.

### Apolipoprotein(a) isoforms

Plasma samples collected from the original YFS participants (Generation 1) during the year 2001 follow-up were stored at –70°C, and assessed in 2015 by western blot to determine apolipoprotein(a) isoforms^16^. All Western blots were inspected by the same researcher. In heterozygous individuals, isoform 1 denotes the smaller (fewer KIV repeats than isoform 2) and isoform 2 the larger apo(a) isoform (more KIV repeats than isoform 1) observed on the Western blot. The relative contribution of isoform 1 to the total blot signal was quantified visually, providing a semiquantitative ranking of the relative expression levels. This defined the dominant isoform, which contributes 50% or more to the Western blot signal, and the nondominant isoform.

### Covariates used in multivariable models

Details of the methods used in the 3-generational YFS have been published^11^. In the present study, in addition to age and sex, the fully adjusted multivariable models included apolipoprotein B (apoB), systolic blood pressure, body mass index (BMI) and smoking as covariates. Serum apoB concentration was determined immunoturbidimetrically with system reagents (Apolipoprotein B, Thermo Fisher Scientific, Finland) on an Indiko Plus analyzer (Thermo Fisher Scientific, Finland). Blood pressure was measured using an automatic Omron HBP-1300 blood pressure measurement device (Omron Corporation, Kyoto, Japan). Because lipid-lowering and blood pressure therapy reduces circulating apoB concentration and lower systolic blood pressure, we created corrected variables intended to approximate the pre-treatment level. For participants reporting statin use, we divided the measured apoB concentration by 0.65. For participants reporting blood pressure medication use, we added 10 mmHg to the measured systolic blood pressure value. This approach follows the common epidemiologic practice of rescaling lipid-related biomarkers in treated individuals by an assumed treatment effect to reduce attenuation bias when modeling associations with outcomes^17,18^. Height and weight were measured. BMI was calculated using the formula: weight [kg]/(height [m])^2^. Smoking habits were inquired with the use of questionnaires.

### Statistical methods

Lp(a) concentrations were analyzed both as particle concentration (nmol/L) and mass concentration (mg/dL). Distributions were right-skewed, therefore, rank-based inverse-normal transformation was used to approximate normality for regression analyses. In addition, Lp(a) was modeled as age- and sex-adjusted quintiles, as well as according to clinically relevant cut-points (≥75 and ≥125 nmol/L for molar units; ≥30 and ≥50 mg/dL for mass units)^7^.

Lp(a) concentrations below the corresponding lower limit of quantification (3.1 mg/dL for mass, and 7.0 nmol/L for molar) were set to the midpoint between 0 and the respective lower limit of quantification (*i.e*. 1.55 mg/dL for mass and 3.5 nmol/L for molar). Because a substantial proportion of participants had Lp(a) values below the assay detection limits (31.9% for molar Lp(a) and 24.3% for Lp(a) mass), these observations shared the same assigned value. As a result, a large number of tied values occurred at the lower end of the distribution, making it impossible to divide the sample into equally sized quintiles. The lowest category contained all individuals with values at the detection limit, and the remaining participants were divided into the higher categories according to the ranked distribution. The association between mass- and molar-based Lp(a) was evaluated using Spearman’s correlation. To assess potential nonlinearity, the functional form of the association was examined using locally weighted scatterplot smoothing (LOESS), a nonparametric method that fits local regressions to the data without assuming a predefined functional form^19^.

Associations between the *LPA* genetic risk score (*LPA* GRS) and Lp(a) concentrations were evaluated using Spearman’s correlation coefficients for molar (nmol/L) and mass (mg/dL) measurements separately. To formally compare the strength of these associations, partial Spearman’s correlations between the *LPA* polygenic risk score and molar and mass Lp(a), adjusted for age and sex, were compared using bootstrap resampling. Because the two coefficients were obtained from the same individuals, the comparison was based on the bootstrap distribution of their within-sample difference over 2000 resamples drawn with replacement. Partial Spearman’s correlations were computed by rank-transforming the variables, residualizing ranked values for age and sex, and correlating the residuals. Bootstrap percentile confidence intervals and a two-sided empirical p value were calculated for the difference between the two coefficients.

To examine structural alignment with apo(a) isoform phenotype, analyses were based on the dominantly expressed apo(a) isoform size and expressed as the number of kringle IV repeats. We specifically tested whether discordance between mass- and molar-based Lp(a) measurements varied according to apo(a) isoform size, and analyzed the ratio of molar to mass Lp(a) in relation to isoform size. Associations with continuous isoform size were evaluated using Spearman’s correlation. In addition, isoform size was categorized into quartiles, and differences in the molar-to-mass ratio across groups were assessed using the Kruskal–Wallis test.

Based on known structural variation in apo(a) isoforms, extreme discordance between mass and molar Lp(a) values is unlikely to be biologically plausible. We therefore performed sensitivity analyses excluding individuals with extreme discordance between molar- and mass-based Lp(a) measurements. This was done by evaluating deviation from the expected relation between the two assays on the original measurement scale. Specifically, log-transformed molar Lp(a) was regressed on log-transformed mass Lp(a), and studentized residuals were calculated. Individuals with an absolute studentized residual >3 (N=120, 1.9%) were classified as having non-physiological discordance and were excluded in sensitivity analyses. This outlier definition was used only for sensitivity analyses, the primary regression analyses used inverse-normal transformed Lp(a) values.

Associations between Lp(a) and atherosclerotic outcomes were examined using logistic regression models. The primary outcome was prevalent CAD, and secondary analyses used a composite cardiovascular disease (CVD) endpoint. Sequential models were fitted: (1) age- and sex-adjusted models; and (2) fully adjusted models including age, sex, apoB, systolic blood pressure, smoking status, and BMI. For quintile-based analyses, the lowest quintile served as the reference category, and global Wald χ² tests were used to evaluate overall association across categories.

Effect estimates are presented as odds ratios (ORs) with 95% confidence intervals (CIs). Model discrimination was assessed using the c-statistic (area under the receiver operating characteristic curve). Model fit was compared using Akaike’s Information Criterion (AIC) and −2 log-likelihood values.

All analyses were performed using SAS version 9.4 (SAS Institute, Cary, NC).

## Results

Lp(a) mass and molar concentrations across age groups are shown in Table 1. Mean Lp(a) levels increased progressively with age for both mass (from 12.8 to 17.2 mg/dL) and molar measurements (from 27.2 to 41.2 nmol/L), with similar age-related shifts observed across the 50th, 75th, and 90th percentiles.

**Table 1.** Age-specific distribution and correlation of Lp(a) measured in mass (mg/dL) and molar (nmol/L) units.

| Age group | N | Lp(a) mass mg/dL |  |  |  | Lp(a) molar nmol/L |  |  |  | Spearman's<br>r |
| --- | --- | --- | --- | --- | --- | --- | --- | --- | --- | --- |
|  |  | Mean | 50<sup>th</sup> | 75<sup>th</sup> | 90<sup>th</sup> | Mean | 50<sup>th</sup> | 75<sup>th</sup> | 90<sup>th</sup> |  |
| 3–17 | 884 | 12.8 | 7.5 | 16.8 | 32.4 | 27.2 | 10.8 | 28.6 | 79.2 | 0.955 |
| 18–29 | 1,202 | 12.7 | 6.8 | 16.8 | 32.2 | 27.8 | 9.4 | 28.6 | 83 | 0.946 |
| 30–49 | 1,197 | 13.9 | 7.7 | 17.6 | 36.6 | 32.0 | 11.8 | 33.7 | 100.1 | 0.950 |
| 50–69 | 1,456 | 15.6 | 8.8 | 20.6 | 39.2 | 37.6 | 14.8 | 41.4 | 109 | 0.951 |
| 70+ | 1,443 | 17.2 | 9.7 | 22.9 | 44.6 | 41.2 | 15.9 | 45.3 | 129.9 | 0.950 |
| All | 6,182 | 14.7 | 8.2 | 19.5 | 37.9 | 34.0 | 12.5 | 37.2 | 101.7 | 0.951 |

Spearman's r indicates rank-order correlation between mass and molar Lp(a) measurements across age-groups.

Molar and mass concentrations of Lp(a) were highly correlated (Spearman’s r=0.951, p<0.0001). The correlation was consistently high across all age groups (r=0.946–0.955), indicating strong agreement between the two measurements throughout the age spectrum. Spearman’s correlations with age were weak for both measures, but highly significant (molar r=0.117; mass r=0.088; both p<0.0001). These associations remained similar after excluding individuals having non-physiological discordance between molar and mass measurements: molar vs. mass r=0.967, molar vs. age r=0.115, mass vs. age r=0.091.

### Analysis of Lp(a) molar-to-mass concentration ratios

Visual inspection (Figure 1) of the scatterplot showed that the relationship between molar and mass based Lp(a) values was approximately linear, with increasing dispersion at higher concentrations. The LOESS curve closely followed the linear regression line across most of the distribution, with only minor deviation at higher Lp(a) concentrations where variability increased. To examine in detail whether the relation between molar and mass Lp(a) measurements remained constant across clinically relevant concentration ranges, we analyzed the molar-to-mass Lp(a) ratio according to established Lp(a) cut-points. The molar-to-mass ratio increased across both molar-based and mass-based Lp(a) categories (Table 2). In nonparametric analyses, testing both overall differences and trends across groups, these between-group differences were significant for both the molar-defined categories (<75, 75–125, and >125 nmol/L; Kruskal–Wallis χ²=1723.2, df=2, p<0.0001) and the mass-defined categories (<30, 30–50, and >50 mg/dL; χ²=1506.4, df=2, p<0.0001). Table 2 includes only individuals with measurable Lp(a) values, but the overall results were unchanged when all participants were included.

**Figure 1.**
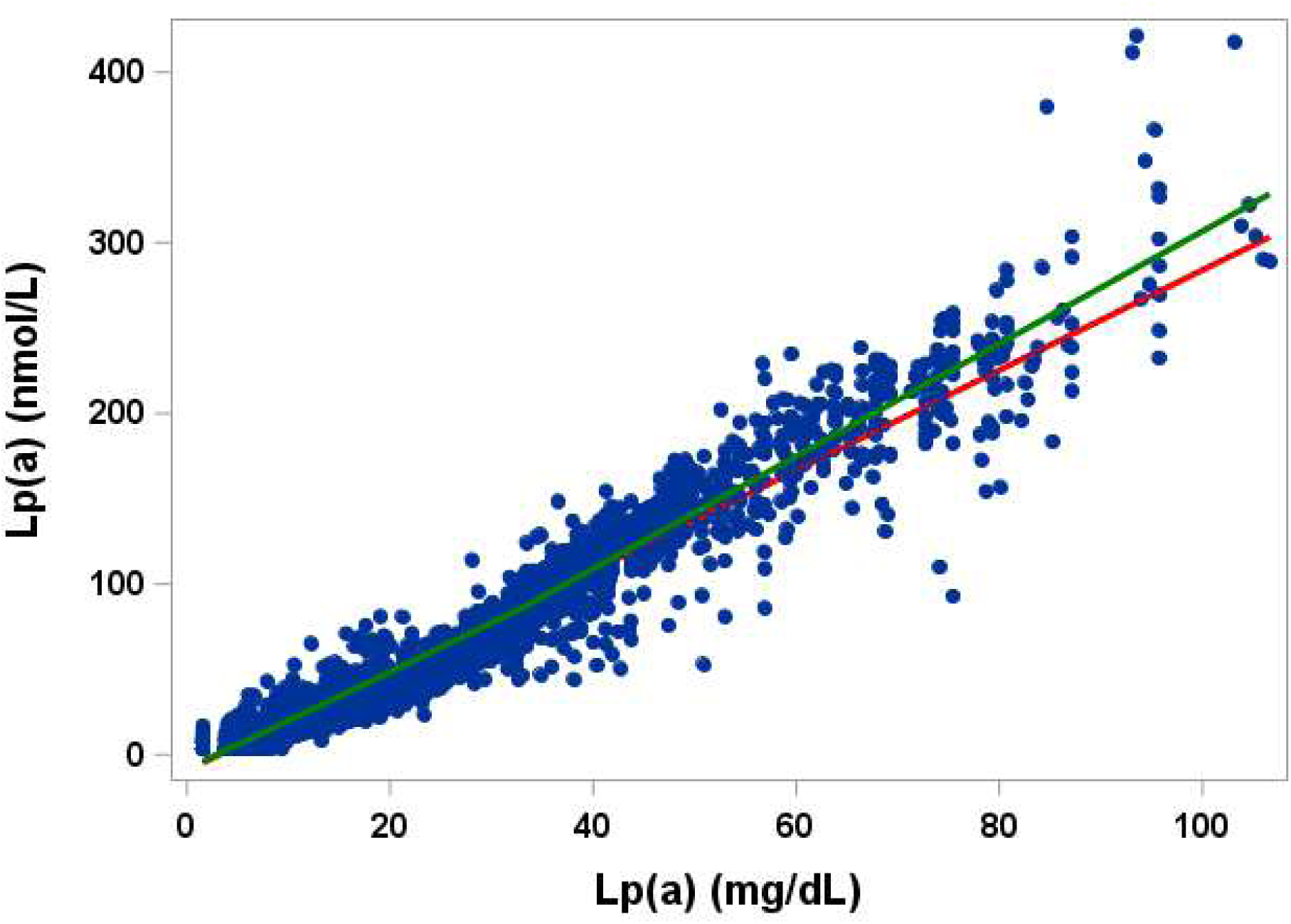
Scatterplot of Lp(a) mass concentration (mg/dL; x-axis) and molar particle concentration (nmol/L; y-axis). The red line represents the fitted linear regression line, and the green line represents the locally weighted scatterplot smoothing (LOESS) curve. The analysis includes 6,062 observations after excluding 120 individuals with non-physiological discordance between mass-and molar-based Lp(a) values.

**Table 2.** Molar-to-mass Lp(a) ratios in relation to clinical Lp(a) cut-points.

| Lp(a) measure | N | Mean ratio | Median ratio |
| --- | --- | --- | --- |
| **Molar (nmol/L)** |  |  |  |
| <75 | 3,228 | 1.75 | 1.61 |
| 75-125 | 401 | 2.70 | 2.68 |
| >125 | 458 | 3.02 | 3.04 |
| P-value (overall) |  |  | <0.0001 |
| P-value (trend) |  |  | <0.0001 |
| **Mass (mg/dL)** |  |  |  |
| <30 | 3,178 | 1.75 | 1.61 |
| 30-50 | 583 | 2.71 | 2.74 |
| >50 | 326 | 2.93 | 2.95 |
| P-value (overall) |  |  | <0.0001 |
| P-value (trend) |  |  | <0.0001 |

### Relations with *LPA* GRS and Lp(a) isoform phenotype

Because genetic variation at the *LPA* locus is a major determinant of Lp(a) levels, we examined whether molar- and mass-based measurements differed in their correlation with the *LPA* GRS, available for 5,680 individuals. The age and sex adjusted partial Spearman’s correlation was nearly identical for molar Lp(a) (r=0.681) than for mass Lp(a) (r=0.674), with both correlations highly significant (p<0.0001). In bootstrap analyses, the difference between partial correlation coefficients was 0.0079 (95% CI 0.0013 to 0.0146; p=0.021), indicating somewhat stronger genetic alignment for molar measurement. The difference between correlation coefficients slightly increased in sensitivity analyses after excluding individuals with non-physiological discordance between molar and mass Lp(a) measurements, and those with values below the assay detection limits: 0.0123 (95% CI 0.0053 to 0.0123; p=0.0005).

To examine whether molar- and mass-based measurements differed in their correlation with the apo(a) size heterogeneity, we analyzed the apo(a) isoform phenotype data that was available for 1,523 individuals. The age and sex adjusted partial Spearman’s correlation was r=-0.535 (p<0.0001) for molar Lp(a) and r=-0.541 (p<0.0001) for mass Lp(a), without evidence of dissimilarity in the bootstrap analyses (p=0.32).

### Associations of the molar-to-mass ratio with *LPA* GRS and apo(a) isoform size

To examine whether the discordance between molar- and mass-based Lp(a) measurements was related to genetic and structural determinants beyond absolute Lp(a) concentration, we analyzed associations of the molar-to-mass ratio with the *LPA* GRS and apo(a) isoform size, with additional adjustment for either molar or mass Lp(a) concentration. The molar-to-mass ratio was directly correlated with the *LPA* GRS (N=5,680; Spearman’s r=0.241, p<0.0001) and inversely correlated with dominant apo(a) isoform size (N=1,523; Spearman’s r=-0.233, p<0.0001). In partial Spearman’s analyses, the association between the molar-to-mass ratio and *LPA* GRS remained significant after adjustment for mass Lp(a) (N=5,680; partial r=0.197, p<0.0001), but not after adjustment for molar Lp(a) (N=5,680; r=0.010, p=0.47). Similarly, for apo(a) isoform size, the inverse association with the molar-to-mass ratio remained significant after adjustment for mass Lp(a) (N=1,523; r=-0.173, p<0.0001), but not after adjustment for molar Lp(a) (r=-0.036, p=0.16). The results of correlation analyses remained essentially similar after excluding individuals with non-physiological discordance between molar and mass Lp(a) measurements, and those with values below the assay detection limits.

### Associations with prevalent CAD

The statistical analyses were restricted to the population aged between 40 and 92 years, as prevalent CAD was very rare in individuals under the age of 40 years (only four cases). In age- and sex-adjusted models, 3,571 individuals were included, of whom 620 (17.4%) had prevalent CAD (Table 3). Both molar- and mass-based Lp(a) measures showed direct associations with prevalent CAD across multiple modeling strategies.

**Table 3.** Associations of Lp(a) with prevalent CAD events.

| Lp(a) Measure | Quintile | Odds Ratio | 95% CI | p-value |
| --- | --- | --- | --- | --- |
| **Models adjusted for age and sex** |  |  |  |  |
| **Molar (nmol/L)** | Q1 | 1 | *Reference* | — |
|  | Q2 | 0.92 | 0.60-1.41 | 0.69 |
|  | Q3 | 0.93 | 0.69-1.25 | 0.64 |
|  | Q4 | 1.44 | 1.09-1.90 | 0.011 |
|  | Q5 | 1.57 | 1.20-2.06 | 0.001 |
| **Mass (mg/dL)** | Q1 | 1 | *Reference* | — |
|  | Q2 | 1.05 | 0.75-1.48 | 0.76 |
|  | Q3 | 1.10 | 0.79-1.53 | 0.57 |
|  | Q4 | 1.48 | 1.08-2.03 | 0.014 |
|  | Q5 | 1.56 | 1.15-2.13 | 0.0049 |
| **Model specification** |  |  |  |  |
| **Molar (nmol/L)** | Lp(a) (continuous) | 1.21 | 1.10-1.34 | 0.0002 |
|  | ≥75 vs <75 | 1.52 | 1.19-1.94 | 0.0009 |
|  | ≥125 vs <125 | 1.54 | 1.14-2.08 | 0.005 |
| **Mass (mg/dL)** | Lp(a) (continuous) | 1.22 | 1.10-1.34 | 0.0001 |
|  | ≥30 vs <30 | 1.43 | 1.12-1.82 | 0.004 |
|  | ≥50 vs <50 | 1.53 | 1.08-2.17 | 0.017 |
| **Fully adjusted models** |  |  |  |  |
| **Molar (nmol/L)** | Q1 | 1 | *Reference* | — |
|  | Q2 | 0.93 | 0.60-1.43 | 0.74 |
|  | Q3 | 0.93 | 0.69-1.26 | 0.63 |
|  | Q4 | 1.43 | 1.08-1.90 | 0.01 |
|  | Q5 | 1.55 | 1.17-2.04 | 0.002 |
| **Mass (mg/dL)** | Q1 | 1 | *Reference* | — |
|  | Q2 | 1.21 | 0.85-1.71 | 0.29 |
|  | Q3 | 1.16 | 0.85-1.60 | 0.35 |
|  | Q4 | 1.63 | 1.20-2.21 | 0.002 |
|  | Q5 | 1.67 | 1.24-2.25 | 0.0008 |
| **Model specification** |  |  |  |  |
| **Molar (nmol/L)** | Lp(a) (continuous) | 1.20 | 1.08-1.33 | 0.0005 |
|  | ≥75 vs <75 | 1.48 | 1.16-1.91 | 0.002 |
|  | ≥125 vs <125 | 1.47 | 1.08-1.98 | 0.014 |
| **Mass (mg/dL)** | Lp(a) (continuous) | 1.22 | 1.10-1.35 | 0.0001 |
|  | ≥30 vs <30 | 1.38 | 1.08-1.77 | 0.01 |
|  | ≥50 vs <50 | 1.42 | 0.99-2.02 | 0.054 |

Odds ratios for continuous models represent the increase in odds per one-unit increase in inverse-normal transformed Lp(a). Fully adjusted models include age, sex, apoB, systolic blood pressure, smoking and BMI, as covariates.

### Age- and sex-adjusted models

When Lp(a) was modeled as quintiles, molar Lp(a) demonstrated a risk gradient, with higher odds of CAD in the upper categories (global Wald χ² 20.8, p=0.0003). Relative to the lowest quintile, the odds ratio (OR) was 1.57 for the highest quintile (p=0.001) (Table 3). Mass-based quintiles were also associated with CAD (global Wald χ² 15.3, p=0.004). Relative to the lowest quintile, the OR was 1.56 for Q5 (p=0.005) (Table 3). Model discrimination was similar between the approaches (c=0.822 for molar vs 0.821 for mass).

In continuous models, both inverse-normal transformed measures were associated with CAD with identical point estimates (OR ∼ 1.2 per 1-unit increase). Using clinical thresholds, elevated molar Lp(a) (≥75 vs <75 nmol/L) was associated with higher odds of CAD (OR 1.52). The corresponding mass threshold (≥30 vs <30 mg/dL) showed a comparable association (OR 1.43). Higher cut-points also identified increased odds, with OR 1.54 for ≥125 vs <125 nmol/L and OR 1.53 for ≥50 vs <50 mg/dL.

### Fully adjusted models

Fully adjusted analyses (using age, sex, apoB, systolic blood pressure, smoking, and BMI as covariates) included 3,568 participants with 618 CAD events (Table 3). In these models, the quintile patterns remained essentially similar for both measures as compared to the age and sex adjusted models (Table 3). Relative to the lowest quintile, the OR was 1.55 for Q5 (p=0.002) for molar Lp(a), and 1.67 for mass Lp(a) (Table 3).

In fully adjusted models for continuous Lp(a), both measures remained independently associated with CAD, with OR 1.20 per unit for molar (p=0.0005) and OR 1.22 for mass (p=0.0001). Using clinical thresholds, molar cut-points remained associated with CAD (≥75 vs <75 nmol/L: OR 1.48, p=0.002; ≥125 vs <125 nmol/L: OR 1.47; p=0.014). Mass thresholds were also associated (≥30 vs <30 mg/dL: OR 1.38; p=0.01; ≥50 vs <50 mg/dL: OR 1.42; p=0.054).

### Associations with composite cardiovascular disease outcome

Among 3,904 participants, 3,891–3,898 had complete data for the composite CVD endpoint and covariates (6–13 excluded for missingness). The prevalence of composite CVD was 24.3% (941/3,891).

In fully adjusted logistic regression models, Lp(a) modeled in molar units showed a graded association with composite CVD across quintiles (global Wald χ² for quintiles: 10.7; p=0.03). Compared with the lowest quintile, the odds of composite CVD were higher in Q4 (OR 1.33, 95% CI 1.06–1.70; p=0.02), and Q5 (OR 1.28, 95% CI 1.01–1.62; p=0.04).

Using mass-based quintiles (mg/dL), the overall association with composite CVD remained statistically significant (global Wald χ² for quintiles: 11.7; p=0.02). Relative to the lowest quintile, higher odds were observed for Q4 (OR 1.47, 95% CI 1.14–1.90; p=0.003) and Q5 (OR 1.36, 95% CI 1.06–1.76; p=0.02).

When modeled as inverse-normal transformed continuous variables in fully adjusted models, both molar and mass Lp(a) were associated with composite CVD: molar Lp(a) OR 1.11 (95% CI 1.02–1.22; p=0.02); mass Lp(a) OR 1.14 (95% CI 1.05–1.25; p=0.002) for mass units, with comparable discrimination (c=0.805 vs 0.806, respectively).

Using clinical cut-points, in age and sex adjusted models, ≥75 nmol/L (molar) was associated with higher odds of composite CVD (OR 1.27, 95% CI 1.02–1.58; p=0.03). The alternative molar threshold, ≥125 nmol/L, had an OR of 1.29 (95% CI 0.99–1.69; p=0.06). For mass-based thresholds, ≥30 mg/dL had an OR of 1.24 (95% CI 1.00–1.53; p=0.05), and the alternative mass threshold, ≥50 mg/dL, had an OR of 1.42 (95% CI 1.05–1.94; p=0.03). After full adjustment, these dichotomous associations were somewhat diluted (molar ≥75 nmol/L: OR 1.24, 95% CI 1.00–1.55; p=0.06; molar ≥125 nmol/L: OR 1.23, 95% CI 0.94–1.62; p=0.13; mass ≥30 mg/dL: OR 1.20, 95% CI 0.97–1.49; p=0.09; mass ≥50 mg/dL: OR 1.33, 95% CI 0.97–1.81; p=0.07).

### Discrimination and Model Fit

Across CAD and composite CVD outcomes, discrimination was nearly identical for molar- and mass-based Lp(a) quintile models. For CAD, the c-statistics were 0.828 for molar and 0.827 for mass, with AIC values of 2553.7 and 2556.1 and −2 log likelihood values of 2531.7 and 2534.1, respectively. For composite CVD, the c-statistics were 0.806 for molar and 0.807 for mass, with AIC values of 3420.8 and 3419.8 and −2 log likelihood values of 3398.8 and 3397.8, respectively. Overall, discrimination and global model fit were highly comparable between the two measurement scales.

### Sensitivity analyses

In sensitivity analyses excluding individuals with non-physiological discordance between molar and mass Lp(a) measurements, as well as those with values below the assay detection limits, the effect estimates for the associations of both molar and mass Lp(a) with CAD and composite CVD remained essentially unchanged.

## Discussion

We found in the YFS population that molar- and mass-based Lp(a) measurements were highly correlated and showed comparable associations with prevalent CAD and composite CVD. In both age- and sex-, as well as in fully adjusted analyses of prevalent CAD and composite CVD, both metrics were associated with disease across quintile, continuous, and clinically thresholded models. Across both CAD and composite CVD, discrimination and global model fit were similar between the two measurement scales, indicating no material difference in overall epidemiologic performance. Thus, the present results suggest that mass- and molar-based Lp(a) provide very similar information at the population level for prevalent cardiovascular outcomes. Similarly, in the ODYSSEY OUTCOMES trial, mass- and molar-based immunoassay, and mass spectrometry–based Lp(a) measurements demonstrated nearly identical associations with recurrent CVD events in a secondary prevention setting^10^. Our population-based analyses are broadly consistent with this overall comparability at the epidemiologic level. At the same time, our genetic and isoform results suggest that molar reporting may align somewhat more closely with the underlying biologic determinants of Lp(a), even if this does not translate into major differences in prevalent disease discrimination.

Consistent with previous observations by Tsimikas *et al.*^9^, we found that the relation between molar and mass-based Lp(a) measurements was not constant across the concentration range. LOESS smoothing indicated that the association was approximately linear across most of the distribution, with modest deviation at higher Lp(a) concentrations. Accordingly, the molar-to-mass ratio increased across higher clinically relevant Lp(a) categories, irrespective whether categorized using molar or mass cut-points. These findings indicate that the correspondence between nmol/L and mg/dL is concentration-dependent rather than fixed, supporting the view that a single universal conversion factor between the two units is not appropriate. In addition to underlying biological differences, assay-related factors may contribute to this pattern. For example, calibration procedures at higher Lp(a) concentrations and the use of polyclonal antibodies that recognize repetitive epitopes in apo(a) may introduce concentration-dependent differences between molar and mass measurements, as suggested by Scharnagl *et al*.^20^.

The genetic and isoform analyses provided additional insight into the biologic basis of this discordance. Both molar and mass Lp(a) were directly correlated with the *LPA* GRS and indirectly with the apo(a) isoform size. Notably, the correlation of the *LPA* GRS was modestly but significantly stronger for molar than for mass Lp(a). Similarly, the molar-to-mass Lp(a) ratio was directly associated with the *LPA* GRS and inversely associated with apo(a) isoform size, indicating higher molar relative to mass values at higher genetically determined Lp(a) burden and in smaller isoforms. To determine whether these associations of the molar-to-mass ratio with *LPA* GRS and apo(a) isoform size merely reflected higher or lower absolute Lp(a) levels, we repeated these analyses after separately adjusting for molar and mass Lp(a) concentrations. These associations were no longer significant after adjustment for molar Lp(a) concentration, but remained evident after adjustment for mass Lp(a) concentration. This observation suggests that the genetically and structurally determined component underlying the difference between reporting scales may be more closely captured by molar than by mass measurement. Genetically and structurally determined differences in Lp(a) biology would be expected to align more directly with molar than with mass measurement. *LPA* GRS and apo(a) isoform size are major determinants of circulating Lp(a) particle concentration, while apo(a) isoform size also contributes to variation in particle mass. Accordingly, molar Lp(a) may more closely approximate particle concentration, whereas mass Lp(a) reflects both particle concentration and particle mass.

At the same time, so-called molar Lp(a) assays should not be interpreted as direct particle-counting methods. Contemporary immunoassays, including the Roche assay used here, are polyclonal antibody–based and therefore may still be influenced by apo(a) structure and epitope presentation, as polyclonal antibodies can recognize multiple epitopes, including repetitive domains within apo(a), which may contribute to residual isoform-related differences between assays. Although calibration is designed to reduce isoform-related bias, complete isoform insensitivity is difficult to achieve, and prior studies suggest that residual bias may persist despite calibration, with concentration-dependent and potentially isoform-related differences between assays^20^. Thus, the pattern observed in our data may reflect not only the true structural biology of Lp(a), but also residual assay-related effects. Overall, molar Lp(a) values should therefore be regarded as assay-calibrated estimates of particle concentration rather than strictly structure-independent measures.

Taken together, our results support a nuanced interpretation of molar reporting. On the one hand, mass- and molar-based Lp(a) performed very similarly in epidemiologic models of prevalent CAD and composite CVD. On the other hand, molar reporting showed slightly stronger alignment with the *LPA* GRS, and the ratio analyses suggested that genetically and structurally determined components of assay discordance were more closely captured by molar than by mass Lp(a). Thus, the rationale for molar reporting in this setting is not that it improves disease discrimination, but rather that it may better reflect the biologic dimension of particle concentration while mass reporting remains comparable for clinical epidemiology. Moreover, distinguishing between analytical approaches is important for assay harmonization and traceability, given that traceability alone does not guarantee equivalent results across methods^21^.

Lp(a) concentrations demonstrated a modest increase across age groups. Although Lp(a) levels are strongly genetically determined and largely resistant to lifestyle modification, small and inconsistent effects have been reported for dietary macronutrient composition, hormonal status, as well as certain diseases and medications^1^. Thus, our data support Lp(a) as an inherited biomarker with limited but detectable age dependence.

## Limitations

Several limitations warrant consideration. First, the analyses are based on prevalent cardiovascular events, introducing potential survivor bias and limiting causal inference regarding incident risk, although this does not affect the established causal role of Lp(a)^22^. Second, although nationwide registry linkage provides near-complete outcome ascertainment, misclassification of diagnoses and heterogeneity in event definitions cannot be fully excluded. Third, Lp(a) measurements were obtained using specific immunoassays, and despite their standardization, assay-dependent differences and calibration variability may affect absolute concentrations and comparability with other cohorts or platforms. Fourth, a small number of participants showed substantial discordance between molar and mass measurements. Although such discordance is unlikely to be explained entirely by apo(a) isoform-related differences in particle mass, we cannot determine with certainty whether these observations reflect technical error, calibration differences, or rare biological variation. This uncertainty was addressed by retaining all observations in the primary analyses and evaluating exclusions in sensitivity analyses. Exclusion of these individuals did not materially change the conclusions. Fifth, Lp(a) mass and molar measurements were not performed contemporaneously, with molar concentrations assessed approximately five years after sample collection. The potential impact of differential storage time is not fully established, although prior data suggest modest but measurable assay-dependent decrease over approximately 3 years, with greater decreases observed in individuals with low–molecular-weight apo(a) isoforms, likely reflecting differences in epitope recognition by immunoassays^23^. Such effects would be expected to attenuate Lp(a) concentrations at higher levels and are therefore unlikely to explain the concentration-dependent divergence observed in the present study. Finally, the cohort is ethnically homogeneous and predominantly of Finnish ancestry, which may limit generalizability to populations with different isoform distributions and genetic architectures^24^.

## Conclusion

In this population-based cohort, molar- and mass-based Lp(a) measurements showed comparable associations with prevalent CAD and composite CVD. However, molar Lp(a) showed slightly closer alignment with the genetic and structural determinants of Lp(a), suggesting that particle-based reporting may better reflect the biologic dimension of Lp(a) while preserving similar epidemiologic performance.

## Data Availability

Anonymized data are available upon reasonable request from the YFS research group (https://youngfinnsstudy.utu.fi/).

## Acknowledgements

Generative artificial intelligence (ChatGPT, OpenAI) was used for language editing and structural refinement of the manuscript.

## Sources of funding

The Young Finns Study has been financially supported by the Academy of Finland: grants 356405, 322098, 286284, 134309 (Eye), 126925, 121584, 124282, 129378 (Salve), 117797 (Gendi), and 141071 (Skidi); the Social Insurance Institution of Finland; Competitive State Research Financing of the Expert Responsibility area of Kuopio, Tampere and Turku University Hospitals (grant X51001); Juho Vainio Foundation; Paavo Nurmi Foundation; Finnish Foundation for Cardiovascular Research; Finnish Cultural Foundation; The Sigrid Juselius Foundation; Tampere Tuberculosis Foundation; Emil Aaltonen Foundation; Yrjö Jahnsson Foundation; Signe and Ane Gyllenberg Foundation; Diabetes Research Foundation of Finnish Diabetes Association; EU Horizon 2020 (grant 755320 for TAXINOMISIS and grant 848146 for To Aition); European Research Council (grant 742927 for MULTIEPIGEN project); Tampere University Hospital Supporting Foundation; Finnish Society of Clinical Chemistry; the Cancer Foundation Finland; pBETTER4U_EU (Preventing obesity through Biologically and bEhaviorally Tailored inTERventions for you; project number: 101080117); CVDLink (EU grant nro. 101137278) and the Jane and Aatos Erkko Foundation.

## Disclosures

The authors report no conflicts of interest.

## Nonstandard Abbreviations and Acronyms

apo(a): apolipoprotein (a)
KIV-2: kringle IV type 2
YFS: Young Finns Study
LPA: GRS LPA genetic risk score
LMW: Low molecular weight

## Notes

### Competing Interest Statement

The authors have declared no competing interest.

### Clinical Trial

This is not a clinical trial.

### Author Declarations

Regional Medical Research Ethics Committee of the Wellbeing Services County of Southwest Finland

## References

1. Kronenberg F, Mora S, Stroes ESG, Ference BA, Arsenault BJ, Berglund L, Dweck MR, Koschinsky M, Lambert G, Mach F, et al. Lipoprotein(a) in atherosclerotic cardiovascular disease and aortic stenosis: a European Atherosclerosis Society consensus statement. Eur. Heart J. 2022;43:3925–3946.

2. Raitakari O, Kivelä A, Pahkala K, Rovio S, Mykkänen J, Ahola-Olli A, Loo BM, Lyytikäinen LP, Lehtimäki T, Kähönen M, et al. Long-term tracking and population characteristics of lipoprotein (a) in the Cardiovascular Risk in Young Finns Study. Atherosclerosis. 2022;356:18–27.

3. Raitakari O, Kartiosuo N, Pahkala K, Hutri-Kähönen N, Bazzano LA, Chen W, Urbina EM, Jacobs DR, Sinaiko A, Steinberger J, et al. Lipoprotein(a) in Youth and Prediction of Major Cardiovascular Outcomes in Adulthood. Circulation. 2023;147:23–31.

4. Kronenberg F, Utermann G. Lipoprotein(a): Resurrected by genetics. J. Intern. Med. 2013;273:6–30.

5. Berglund L, Ramakrishnan R. Lipoprotein(a): an elusive cardiovascular risk factor. Arterioscler. Thromb. Vasc. Biol. 2004;24:2219–2226.

6. Crouse JR, Parks JS, Schey HM, Kahl FR. Studies of low density lipoprotein molecular weight in human beings with coronary artery disease. J. Lipid Res. 1985;26:566–574.

7. Kamstrup PR, Neely RDG, Nissen S, Landmesser U, Haghikia A, Costa-Scharplatz M, Abbas C, Nordestgaard BG. Lipoprotein(a) and cardiovascular disease: sifting the evidence to guide future research. Eur. J. Prev. Cardiol. 2024;31:903–914.

8. Koschinsky ML, Bajaj A, Boffa MB, Dixon DL, Ferdinand KC, Gidding SS, Gill EA, Jacobson TA, Michos ED, Safarova MS, et al. A focused update to the 2019 NLA scientific statement on use of lipoprotein(a) in clinical practice. J. Clin. Lipidol. 2024;18:e308–e319.

9. Tsimikas S, Fazio S, Viney NJ, Xia S, Witztum JL, Marcovina SM. Relationship of lipoprotein(a) molar concentrations and mass according to lipoprotein(a) thresholds and apolipoprotein(a) isoform size. J. Clin. Lipidol. 2018;12:1313–1323.

10. Szarek M, Reijnders E, Jukema JW, Bhatt DL, Bittner VA, Diaz R, Fazio S, Garon G, Goodman SG, Harrington RA, et al. Relating Lipoprotein(a) Concentrations to Cardiovascular Event Risk After Acute Coronary Syndrome: A Comparison of 3 Tests. Circulation. 2024;149:192–203.

11. Pahkala K, Rovio S, Kartiosuo N, Auranen K, Bourgery M, Elovainio M, Fogelholm M, Haapala J, Hirvensalo M, Hutri N, et al. Cohort Profile Update: Expanding the Cardiovascular Risk in Young Finns Study into a multigenerational cohort. Int. J. Epidemiol. 2026;55:dyaf206.

12. Raitakari OT, Juonala M, Rönnemaa T, Keltikangas-Järvinen L, Räsänen L, Pietikäinen M, Hutri-Kähönen N, Taittonen L, Jokinen E, Marniemi J, et al. Cohort profile: The cardiovascular risk in young Finns study. Int. J. Epidemiol. 2008;37:1220–1226.

13. Jacobs DR, Woo JG, Sinaiko AR, Daniels SR, Ikonen J, Juonala M, Kartiosuo N, Lehtimäki T, Magnussen CG, Viikari JSA, et al. Cardiovascular Risk Factors in Childhood and Adult Cardiovascular Events. N. Engl. J. Med. 2022;386:1877–1888.

14. Das S, Forer L, Schönherr S, Sidore C, Locke AE, Kwong A, Vrieze SI, Chew EY, Levy S, McGue M, et al. Next-generation genotype imputation service and methods. Nat. Genet. 2016;48:1284–1287.

15. Trinder M, Paruchuri K, Haidermota S, Bernardo R, Zekavat SM, Gilliland T, Januzzi J, Natarajan P. Repeat Measures of Lipoprotein(a) Molar Concentration and Cardiovascular Risk. J. Am. Coll. Cardiol. 2022;79:617–628.

16. Kronenberg F, Kuen E, Ritz E, Junker R, König P, Kraatz G, Lhotta K, Mann JFE, Müller GA, Neyer U, et al. Lipoprotein(a) serum concentrations and apolipoprotein(a) phenotypes in mild and moderate renal failure. J. Am. Soc. Nephrol. 2000;11:105–115.

17. Peloso GM, Auer PL, Bis JC, Voorman A, Morrison AC, Stitziel NO, Brody JA, Khetarpal SA, Crosby JR, Fornage M, et al. Association of low-frequency and rare coding-sequence variants with blood lipids and coronary heart disease in 56,000 whites and blacks. Am. J. Hum. Genet. 2014;94:223–232.

18. Surendran P, Feofanova E V., Lahrouchi N, Ntalla I, Karthikeyan S, Cook J, Chen L, Mifsud B, Yao C, Kraja AT, et al. Discovery of rare variants associated with blood pressure regulation through meta-analysis of 1.3 million individuals. Nat. Genet. 2020;52:1314–1332.

19. Cleveland WS. Robust Locally Weighted Regression and Smoothing Scatterplots. J. Am. Stat. Assoc. 1979;74:829.

20. Scharnagl H, Stojakovic T, Dieplinger B, Dieplinger H, Erhart G, Kostner GM, Herrmann M, März W, Grammer TB. Comparison of lipoprotein (a) serum concentrations measured by six commercially available immunoassays. Atherosclerosis. 2019;289:206–213.

21. Badrick T, Beasley-Green A, Cobbaert CM, Delatour V, Deprez L, Jones GRD, Josephs RD, Kessler A, Liu Q, Maniguet S, et al. Result harmonization in medical laboratories: accomplishments and challenges. Clin. Chem. Lab. Med. 2026;

22. Kronenberg F. Human Genetics and the Causal Role of Lipoprotein(a) for Various Diseases. Cardiovasc. Drugs Ther. 2016;30:87–100.

23. Kronenberg F, Trenkwalder E, Dieplinger H, Utermann G. Lipoprotein(a) in stored plasma samples and the ravages of time. Why epidemiological studies might fail. Arterioscler. Thromb. Vasc. Biol. 1996;16:1568–1572.

24. Erhart G, Lamina C, Lehtimäki T, Marques-Vidal P, Kähönen M, Vollenweider P, Raitakari OT, Waeber G, Thorand B, Strauch K, et al. Genetic factors explain a major fraction of the 50% lower lipoprotein(a) concentrations in Finns. Arterioscler. Thromb. Vasc. Biol. 2018;38:1230–1241.

